# Predictive power of youth diabetes screening guidelines

**DOI:** 10.64898/2025.12.01.25341261

**Authors:** Sarah M. Halvorson-Fried, Nita Vangeepuram, Negar Golestani, Bian Liu

**Affiliations:** Division of General Internal Medicine, Icahn School of Medicine at Mount Sinai; Tisch Cancer Institute, Icahn School of Medicine at Mount Sinai; Department of Pediatrics, Icahn School of Medicine at Mount Sinai; Department of Population Health Science and Policy, Icahn School of Medicine at Mount Sinai; Institute for Health Equity Research, Icahn School of Medicine at Mount Sinai; Department of Artificial Intelligence and Human Health, Icahn School of Medicine at Mount Sinai; Department of Environmental Medicine, Icahn School of Medicine at Mount Sinai

## Abstract

**Introduction:** Early-onset type 2 diabetes (EOD), defined as type 2 diabetes diagnosed before age 45, is a growing public health problem. Adolescence is a critical period for prevention interventions because related health behaviors and biological processes are established during this time. Early screening for type 2 diabetes risk including prediabetes (preDM) could identify youth most in need of intervention but unlike for adults, no questionnaire-based youth risk screeners exist. In this study, we assessed the ability of adapted American Diabetes Association (ADA) screening guidelines in adolescence to predict EOD and prediabetes (preDM/EOD) in adulthood.

**Methods:** We used data from the National Longitudinal Study of Adolescent to Adult Health (Add Health). We applied ADA guidelines (modified to align with available data) in Waves I and II, when participants were 12-19, and assessed ability to predict preDM/EOD in Waves IV and V, when participants were 24-43. We then calculated screening performance measures with (N=13,530) and without (N=14,540) accounting for the survey design.

**Results:** In weighted analyses, 40% of participants (5,383) had preDM/EOD based on biomarkers at Waves IV-V. Of these participants, 1,272 were considered at risk according to the ADA screening criteria in Waves I-II (sensitivity=23.6%). Of 8,147 participants without preDM/EOD, 7,218 were not considered at risk in Wave I-II (specificity=88.6%). The screening guidelines had a positive predictive value (PPV) of 57.8% and a negative predictive value (NPV) of 63.7%. The F+ and F- measures were 33.5% and 74.1%, respectively. Unweighted analyses produced similar results.

**Conclusions:** Adapted ADA youth screening guidelines demonstrated poor ability to predict preDM/EOD before age 45. Although specificity was high, sensitivity was low: 76% of participants who developed preDM/EOD would not have been identified as at risk in adolescence using the adapted ADA guidelines. Given the high prevalence of preDM/EOD, there is a need for better screening tools to identify youth at risk and better target diabetes prevention interventions.

## Introduction

Early-onset type 2 diabetes (EOD), defined as type 2 diabetes diagnosed before age 45, is a growing public health problem.^1^ Approximately 6 million adults aged 18-44 have diabetes in the US, including 2 million with undiagnosed diabetes.^2^ Nearly 33 million adults aged 18-44 have prediabetes (preDM).^2^ EOD is particularly harmful because it leads to earlier onset of complications compared with type 2 diabetes in older adulthood.^1^ Adolescence is a critical period for prevention because related health behaviors^3^ and biological processes^1^ are established during this time.

Screening youth for diabetes risk could help better target prevention efforts, but unlike for adults, no questionnaire-based youth risk screeners exist. We examined adapted American Diabetes Association (ADA) clinical guidelines for identifying adolescents (ages 12-19)^4^ with preDM/EOD and found they had limited performance.^5^ In this study, we assessed the ability of the adapted ADA screening guidelines in adolescence to predict preDM/EOD in adulthood using nationally representative, longitudinal data.

## Methods

We used data from the National Longitudinal Study of Adolescent to Adult Health (Add Health).^6^ We applied ADA guidelines (modified to align with available data, Table 1) in Waves I and II, when participants were 12-19, and assessed ability to predict preDM/EOD in Waves IV and V, when participants were 24-43. We defined preDM/EOD as HbA1c ≥5.7%, fasting glucose ≥100mg/dL, and/or non-fasting glucose ≥140 mg/dL. We then calculated screening performance measures.

**Table 1.** ADA risk factors and Add Health variables used for risk factors.

| **ADA preDM/EOD risk criteria**<br>**for children**<br>**(at-risk if overweight plus 1+**<br>**additional factors)** | **Add Health**<br>**variables used** | **Prevalence**<br>**[No. (%)]** |  |
| --- | --- | --- | --- |
|  |  | **Weighted**<br>**(N = 13,530)** | **Unweighted**<br>**(N = 14,540)** |
| Overweight (BMI ≥ 85th percentile for age and sex) | Overweight (BMI ≥ 85th percentile for age and sex) | 4,009 (29.6%) | 4,232 (29.1%) |
| Maternal history of gestational diabetes during the child's gestation | Not available | N/A | N/A |
| Family history of type 2 diabetes in first- or second-degree relative | Diagnosis of diabetes in biological mother or father or diabetes/high blood sugar for grandparent, aunt, or uncle | 2,467 (18.2%) | 1,653 (11.4%) |
| Race/ethnicity (Native American, African American, Latino, Asian American, Pacific Islander) | Race/ethnicity (NH Native American, Black/African American, Hispanic/Latino, NH Asian/Pacific Islander) | 4,229 (31.3%) | 6,438 (44.3%) |
| Signs of insulin resistance or conditions associated with insulin resistance (hypertension, dyslipidemia, acanthosis nigricans, polycystic ovary syndrome, or small-for-gestational-age birth weight) | Self-report of hypertension or dyslipidemia before age 20 (reported at later wave) or birth weight < 2500g | 1,205 (8.9%) | 1,464 (10.1%) |

*Notes.* ADA = American Diabetes Association. PreDM = Prediabetes. EOD = Early onset diabetes. NH = Non-Hispanic. Weighted analyses use Wave IV survey weights and account for Add Health's complex survey design.

We ran analyses with and without accounting for the Add Health survey design using strata, clustering, and weights.^6^ For unweighted analyses, we limited the sample to participants who had non-missing risk values in Wave I-II and at least one non-missing biomarker in Waves IV-V (N=14,540). For weighted analyses, we used Wave IV survey weights because the majority of participants (n=9,818, 68%) were missing biomarker data for Wave V. We excluded 1,010 participants missing survey weights (N=13,530).

## Results

In weighted analyses, 40% of participants (5,383) had preDM/EOD based on biomarkers at Waves IV-V. Of these participants, 1,272 were considered at risk according to the ADA screening criteria in Waves I-II (sensitivity=23.6%, Table 2). Of 8,147 participants without preDM/EOD, 7,218 were not considered at risk in Wave I-II (specificity=88.6%). The screening guidelines had a positive predictive value (PPV) of 57.8% and a negative predictive value (NPV) of 63.7%. The F+ and F-measures were 33.5% and 74.1%, respectively. Unweighted analyses produced similar results (Table 2).

**Table 2.** Performance measures of ADA clinical screening guidelines at Wave I-II (ages 12-19) compared with preDM/EOD at Wave IV-V (ages 24-43).

| Measure | Weighted<br>(N = 13,530) | Unweighted<br>(N = 14,738) |
| --- | --- | --- |
| Screeener +, PreDM/EOD + | 1,272 | 1,570 |
| Screeener -, PreDM/EOD + | 4,111 | 4,465 |
| Screeener +, PreDM/EOD - | 929 | 1,090 |
| Screeener -, PreDM/EOD - | 7,218 | 7,415 |
| Total PreDM/EOD + | 5,383 | 6,035 |
| Total PreDM/EOD - | 8,147 | 8,505 |
| Total Screeener + | 2,201 | 2,660 |
| Total Screeener - | 11,329 | 11,880 |
| Sensitivity | 1,272/5,383 = 23.6% | 1,570/6,035 = 26.0% |
| Specificity | 7,218/8,147 = 88.6% | 7,415/8,505 = 87.2% |
| Positive Predictive Value | 1,272/2,201 = 57.8% | 1,570/2,660 = 59.0% |
| Negative Predictive Value | 7,218/11,329 = 63.7% | 7,415/11,880 = 62.4% |
| F-measure + | $2 * (57.8\% * 23.6\%) / (57.8\% + 23.6\%) = 33.5\%$ | $2 * (59.0\% * 26.0\%) / (59.0\% + 26.0\%) = 36.1\%$ |
| F-measure - | $2 * (63.7\% * 88.6\%) / (63.7\% + 88.6\%) = 74.1\%$ | $2 * (62.4\% * 87.2\%) / (62.4\% + 87.2\%) = 72.7\%$ |

*Notes.* ADA = American Diabetes Association. PreDM = Prediabetes. EOD = Early onset diabetes. Weighted analyses use Wave IV survey weights and account for Add Health's complex survey design.

## Discussion

Adapted ADA youth screening guidelines demonstrated poor ability to predict preDM/EOD before age 45. Although specificity was high, sensitivity was low: 76% of participants who developed preDM/EOD would not have been identified as at risk in adolescence using the adapted ADA guidelines. These findings add to previous work showing poor predictive performance of the ADA guidelines for preDM/EOD among youth.^5^

We were unable to distinguish between type 1 and 2 diabetes; however, since we excluded participants who reported diabetes between ages 12 and 19, we likely captured few, if any, type 1 cases. The ADA screening criteria are not intended to predict preDM/EOD later in life; however, since diabetes risk progresses through the life course, our findings add to an understanding of the screening guidelines’ performance long-term. Strengths include a large, nationally representative longitudinal sample and biomarker-based assessment of preDM/EOD.

Given the high prevalence of preDM/EOD,^2^ there is a need for better screening tools to identify youth at risk and better target diabetes prevention interventions.

## Data Availability

Data used in the present study may be accessed by applying for a restricted-use data contract with the National Longitudinal Study of Adolescent to Adult Health.

https://addhealth.cpc.unc.edu/data/

## Acknowledgments

Research reported in this publication was supported by the National Cancer Institute of the National Institutes of Health under award number T32CA225617. Waves I-V of Add Health were funded by grant P01 HD31921 from the Eunice Kennedy Shriver National Institute of Child Health and Human Development, with cooperative funding from 23 other federal agencies and foundations. Add Health is currently directed by Robert A. Hummer at the University of North Carolina at Chapel Hill. Add Health was designed by J. Richard Udry, Peter S. Bearman, and Kathleen Mullan Harris at the University of North Carolina at Chapel Hill. The Add Health Parent Study/Parents (2015-2017) data collection was funded by a grant from the National Institute on Aging (RO1AG042794) to Duke University, V. Joseph Hotz (PI) and the Carolina Population Center at the University of North Carolina at Chapel Hill, Kathleen Mullan Harris (PI).

## References

1. Wilmot E, Idris I. Early onset type 2 diabetes: risk factors, clinical impact and management. Ther Adv Chronic Dis. 2014;5(6):234-244. doi:10.1177/2040622314548679

2. Centers for Disease Control and Prevention. National Diabetes Statistics Report. May 15, 2024. Accessed October 8, 2025. https://www-cdc-gov.libproxy.lib.unc.edu/diabetes/php/data-research/index.html

3. Lawrence E, Mollborn S, Goode J, Pampel F. Health lifestyles and the transition to adulthood. Socius. 2020;6:2378023120942070. doi:10.1177/2378023120942070

4. Arslanian S, Bacha F, Grey M, Marcus MD, White NH, Zeitler P. Evaluation and management of youth-onset type 2 diabetes: a position statement by the American Diabetes Association. Diabetes Care. 2018;41(12):2648-2668. doi:10.2337/dci18-0052

5. Vangeepuram N, Liu B, Chiu Po-hsiang, Wang L, Pandey G. Predicting youth diabetes risk using NHANES data and machine learning. Sci Rep. 2021;11(1):11212. doi:10.1038/s41598-021-90406-0

6. Harris KM. The National Longitudinal Study of Adolescent to Adult Health (Add Health), Waves I & II, 1994–1996; Wave III, 2001–2002; Wave IV, 2007-2009; Wave V, 2016-2018 [machine-readable data file and documentation]. Published online 2018.

